# Biomarkers for Identification of High-Risk Coronary Artery Plaques in Patients with Suspected Coronary Artery Disease

**DOI:** 10.1101/2023.09.14.23295593

**Authors:** Gitte Pedersen, Jonathan Nørtoft Dahl, Laust Dupont Rasmussen, Anne-Birgitte Garm Blavnsfeldt, Sidsel Hauge Böttcher, Matias Hauge Böttcher, Mette Nyegaard, Louise Nissen, Simon Winther

## Abstract

**Background:** Patients with atherosclerotic plaques containing high-risk features have an increased likelihood of events and a worse prognosis. Whether increased levels of Troponin I (TnI) and C-reactive protein (CRP) are associated with the presence of high-risk coronary atherosclerotic plaques (HRP) is not well described. We assessed the association between 1) TnI and 2) CRP with quantified coronary plaque burden, luminal diameter stenosis, and HRP in patients with low/intermediate pre-test probability of obstructive coronary artery disease (CAD) referred for coronary computed tomography angiography (CCTA).

**Methods:** The CCTA from 1,615 patients were analyzed using a semiautomatic software for coronary artery plaque characterization. Patients with high TnI (>6 ng/L) and high CRP (>2 mg/L) were identified. Associations of TnI and CRP with plaque burden, stenosis (≥50% luminal diameter stenosis on CCTA), and HRP were investigated.

**Results:** TnI and CRP were both positively correlated with total plaque burden (TnI r_s_=0.14, p<0.001; CRP r_s_=0.08, p<0.001). In multivariate logistic regression analyses, high TnI was associated with stenosis (OR 1.43, 95% confidence interval (CI) 1.03-1.99, p=0.034), the presence of HRP (OR 1.79, 95% CI: 1.17– 2.74, p=0.008), and the subtypes of HRP; low attenuation plaque (OR 1.93, 95% CI: 1.24–3.00, p=0.003), and positive remodeling (OR 1.51, 95% CI: 1.07–2.13, p=0.018). For CRP, only stenosis and napkin ring sign correlated significantly.

**Conclusion:** In patients with suspected CAD, TnI and CRP are associated with HRP features. These findings may suggest that inflammatory and particularly ischemic biomarkers might improve early risk stratification and affect patient management. ClinicalTrials.gov identifier: NCT02264717

**CLINICAL PERSPECTIVE:** Using CCTA, our findings direct the focus toward plaque characteristics rather than just overall plaque burden, outlining that the presence of stenosis and specifically HRPs may be more important in CAD risk evaluation than the amount of atherosclerosis alone. Our findings suggest that biomarkers can help identify patients with HRP features, which previously were shown to increase the risk of future events. TnI may have a place in pre-test evaluation of patients with stable chest pain by introducing biomarkers to a pre-test clinical likelihood model, which may pave the way for more accurate risk stratification and, consequently, better-informed clinical decision-making. Still, trials on biomarker-guided diagnostic testing and medical therapy in de novo stable chest pain patients are warranted.

## INTRODUCTION

Coronary artery disease (CAD) is a widespread cause of morbidity and mortality. In patients with suspected CAD, coronary computed tomography angiography (CCTA) is a first-line non-invasive test with high diagnostic accuracy of obstructive disease.^*1*^ In general, patients with de novo symptoms suggestive of obstructive CAD show favorable prognosis,^*2*^ however CCTA is able to identify non-obstructive coronary plaques, plaque burden, and high-risk plaque (HRP) features which predict adverse cardiovascular events.^*3*–*5*^ Positive remodeling, low attenuation plaques, napkin ring sign, and spotty calcifications are examples of HRP features which often serve as culprits in later coronary events.^*3*, *4*, *6*^ Similar to the presence of HRP, cardiovascular risk factors, inflammation, and myocardial ischemia increase event rates.^*7*, *8*^

To improve identification of patients with de novo chest pain and elevated risk of cardiovascular events, several biomarkers have been suggested.^*8*, *9*^ Troponin I (TnI) detects myocardial necrosis and cell death, and C-reactive protein (CRP) low-grade inflammation, both linked to CAD events^*7*, *10*, *11*^ Yet, the relationship between these biomarkers with plaque burden and HRPs is not well described therefore existing findings warrant validation, and clinical thresholds for TnI and CRP to predict coronary characteristics to enable patient-specific risk stratification remain undetermined.^*12*^ In general, usage of circulating biomarkers could potentially improve early risk stratification and patient management of patients with de novo symptoms suggestive of obstructive CAD.

This study investigated if TnI and CRP are associated with plaque burden, stenosis, and HRP on CCTA.

## METHODS

### Patients and study design

This was a sub-study of the Danish study of Non-Invasive testing in Coronary Artery Disease (Dan-NICAD) 1 trial. The study protocol and main results were previously reported.^*13*, *14*^ Patients were enrolled from September 11th 2014 until March 31th 2016 and inclusion and exclusion criteria are described in supplemental table 1. In short, the trial was designed to determine the diagnostic accuracy of different second-line non-invasive tests after CCTA with suspected stenosis using invasive coronary angiography with fractional flow reserve (FFR) as reference for obstructive CAD. Blood was sampled at the day of CCTA, and CCTAs were qualitatively and quantitatively analyzed post hoc at a core lab facility to evaluate plaque burden, stenosis, and HRPs. Patients were excluded if blood samples were missing or CCTA had poor image quality or was not performed.

Patients provided informed consent. The study was approved by The Danish Data Protection Agency and the Central Denmark Regional Committee on Health Research Ethics and registered at ClinicalTrials.gov.

### Data collection

Biochemistry, risk factors, medication, and medical history data were obtained prior to CCTA through interviews by trained nurses and electronic medical records. Blood was sampled on the day of the CCTA.

#### Analyses of High-sensitive C-reactive protein and High-sensitive Troponin I

Lithium-heparin plasma samples were collected from all patients before the administration of X-ray contrast medium and stored at -80°C until time of analysis.^*14*^

High-sensitivity TnI and high-sensitivity CRP were measured with the High-Sensitivity Troponin I (TNIH)-analysis chemiluminescence immunoassay (ADVIA Centaur® XPT, Siemens Healthcare Diagnostics) and turbidimetric assay (CardioPhaseTM High-Sensitivity C-reactive protein (ADVIA® Chemistry XPT, Siemens Healthcare Diagnostics)), respectively. Both commercially available and clinically used. The clinical cut-off for potential myocardial injury was 47 ng/L, while the analytical measuring interval for both assays was 3-35,000 ng/l and 0.2-200 mg/l, respectively.

Analyses were performed in the internationally accredited laboratory at the Department of Biochemistry, Aarhus University Hospital, Aarhus, Denmark (DS/EN ISO/IEC 15189).

#### Grouping of High-sensitive C-reactive protein and High-sensitive Troponin I

Using cut-off values based on limit of detection and previous studies,^*15*^ TnI were divided according to the cut-offs at 3.0 ng/L and 6.0 ng/L stratifying patients into three groups as low (*<3.0), moderate (3.0-6.0); and high (>6.0) ng/L. Similarly,* using cut-off values specified from previous studies, CRP was divided into three groups with cut-off at 1 mg/L and 2 mg/L stratifying patients into three groups as low (***<****1.0); moderate (1.0-2.0); and high (>2.0) mg/L.*^*11*, *16*^

#### CT-acquisition

Scans were performed on a 320-slice volume CT scanner (Aquilion One, Toshiba Medical Systems, Japan) as per the study protocol.^*14*^ Sublingual nitroglycerin and beta-blockade were administered to achieve a heart rate <60 beats/min.

#### Analysis of coronary plaques

Plaque analyses were performed on the acquired CCTAs in a core lab setting using a semiautomatic CCTA plaque quantification and characterization software (QAngio® CT Research Edition, Medis, the Netherland, version 3.1.2.4) The plaque analyses were performed by four trained investigators blinded to patient information, and each reader analyzing one-fourth of the assigned images. An expert reader reviewed the analyses for consistency.

With manual adjustments allowed, the software used for plaque quantification and characterization extracted the coronary artery tree, the coronary lumen, and vessel wall contours, thereby enabling both a qualitative and quantitative characterization of vessel morphology, plaque intensities, and overall plaque burden. Scans with low quality due to artifacts, incomplete CCTA processing or insufficient coronary coverage were excluded.

#### Qualitative plaque assessment

Coronary artery calcium score (CACS) was calculated using the Agaston method. Significant diameter stenosis was defined as a ≥50% luminal diameter stenosis in any vessel segment with at diameter of >2mm at initial visual core lab evaluation.^*17*^ Subsequently, qualitative HRP features were evaluated including 1) low-attenuation plaques towards lumen ≤30 Hounsfield units (HU), 2) positive remodeling index of >1.10 (determined by maximum lesion area/healthy reference vessel area), c) napkin ring sign (central low attenuation core, surrounded by a thin semi-circular plaque of lighter attenuation in the outer contour), and d) soft plaque with spotty calcifications encased in soft plaque (≤3mm length calcification).^*18*, *19*^

A high-risk plaque (HRP) was defined as a plaque with at least two of the above features.^*20*^

#### Quantitative plaque assessment

All vessels with a lumen ≥1.8 mm were analyzed, and coronary volumes were quantified using the "no-gap-method."^*21*^ Plaque burden was evaluated on a per patient-level using the following equation for all validated plaque-intensities; total percentage atheroma volume (PAV) = (total plaque volume mm^*3*^ / vessel volume mm^*3*^) ∗100. A high plaque burden was defined as a total plaque burden above the 75th percentile.

Necrotic core was defined as -30 to 30 HU, fatty fibrous 31 to 130 HU, fibrous 131 to 350 HU, and dense calcium >350 HU. Quantitative plaque analyses were performed only in patients with at least one visual plaque at the qualitative assessment. In patients without any visible plaque, both qualitative and quantitative plaque variables were set to zero.^*21*^

### Statistics

Data distribution was evaluated using histograms, QQ-plots, and Shapiro-Wilk tests. Quantitative data was presented as mean (± standard derivation (SD)) or median [interquartile range (IQR)] according to normality, and categorical variables as n (%).

For normally distributed continuous variables, Student’s t-test for independent samples was used, whereas the Mann-Whitney U-tests for independent samples were used for non-normally distributed variables. In case of more than two groups, the Kruskal-Wallis test was used. Proportions of the dichotomous variables were compared using a χ2 analysis.

Correlations between plaque burden (PAV and CACS) and biomarkers were analyzed using Spearman’s correlation coefficients.

Uni- and multivariate logistic regression analyses were performed on a per patient-level to assess the relationship between PAV, stenosis, HRP, and the individual risk features, with TnI and CRP as independent variables. Multivariate analyses were adjusted for the components from a validated pre-test likelihood score of obstructive CAD and cardiac events including sex, age, symptom characteristics, smoking-status, dyslipidemia, diabetes mellitus, antihypertensive therapy, and family history.^*2*, *22*^ Multicollinearity was checked, and missing data were excluded from the logistic regression analysis. Data on biomarkers was categorized according to pre-specified cut-offs defined above. A p-value <0.05 was considered significant.

Data were analyzed using STATA version 17.0.^*23*^

## RESULTS

### Study population

1615 patients with low and intermediate pre-test probability of obstructive CAD, with complete CCTA analyses and biomarker information, were available for statistical analyses *(figure 1, figure 2)*. Baseline characteristics stratified for presence of HRP are summarized in table 1. Patients with HRP were more often men, older, and had more risk factors and typical chest pain compared to patients without HRP *(supplemental table 2)*. The 75th percentile for total plaque burden was 6.95% plaque volume relative to the vessel volume. In total, 376 patients (23.3%) had stenosis, 181 (11%) had HRP, 157 (9.7%) had low attenuation plaque, 340 (21%) had positive remodeling, 39 (2.4%) had napkin ring sign, and 197 (12.2%) had spotty calcifications.

**Figure 1:**
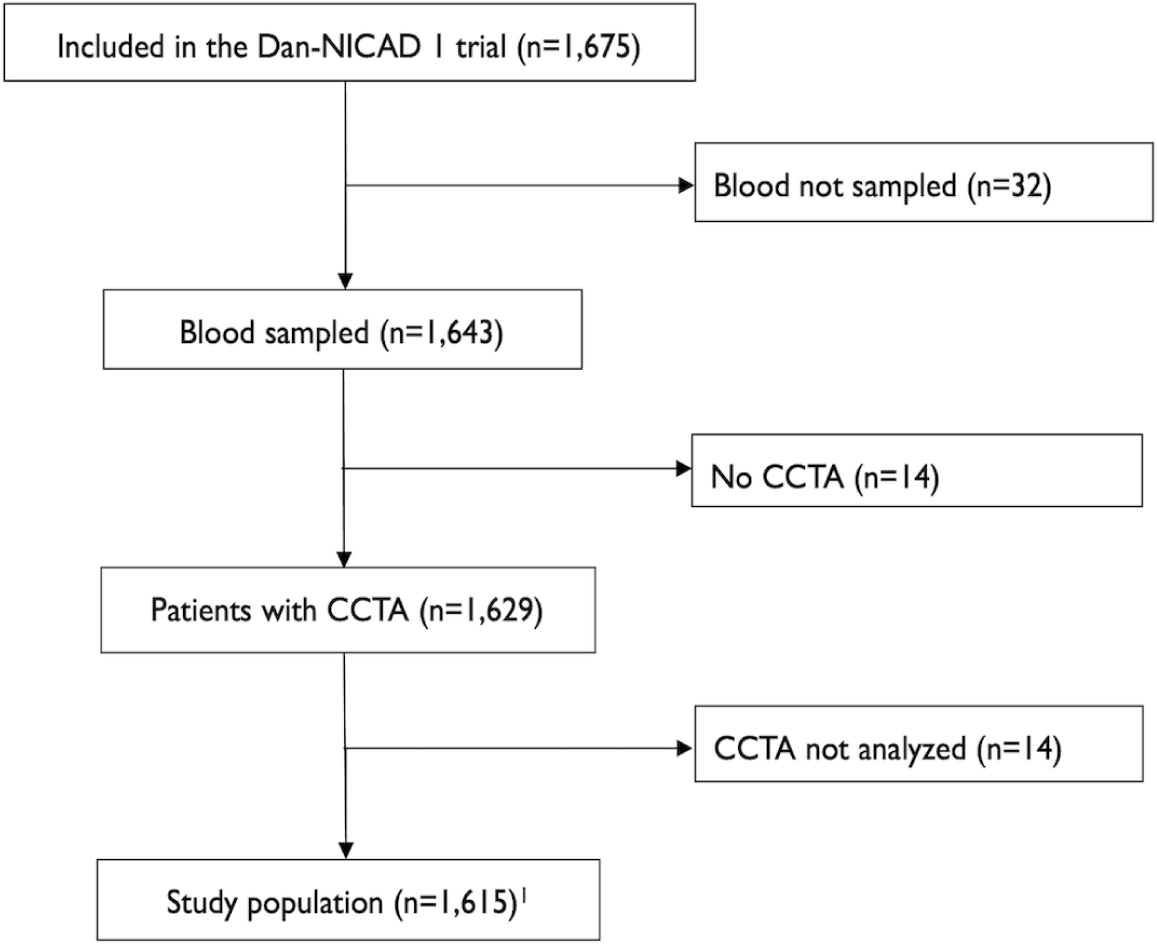
Study flow. ^1^From the 1,675 patients enrolled, 1,615 patients had data both CCTA and CRP. 1,604 patients had data on both CCTA and TnI. CCTA = coronary computed tomography angiography.

**Figure 2:**
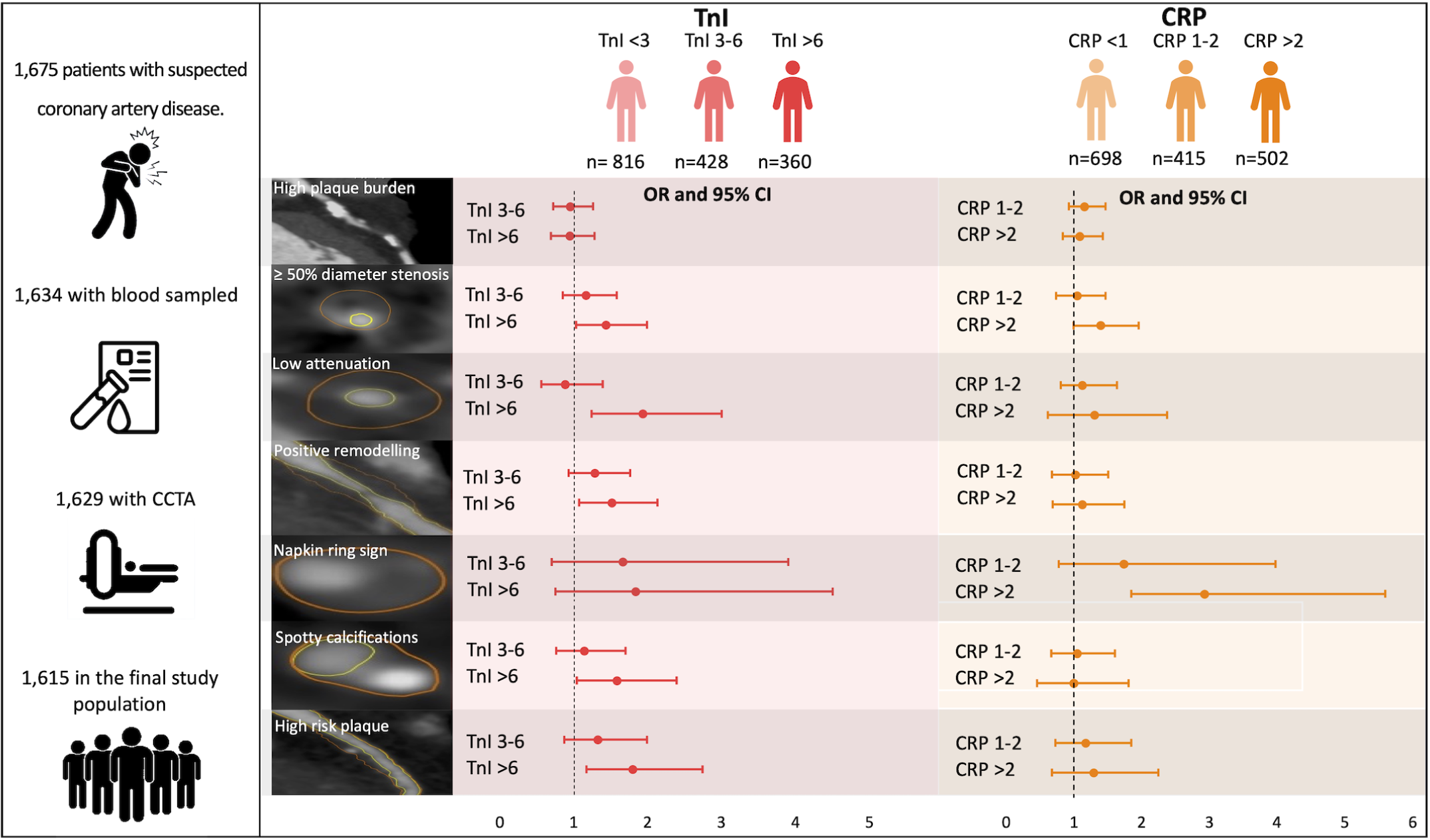
Biomarkers for identification of high plaque burden, significant stenosis and high-risk coronary plaques in patients with suspected coronary artery disease. In multivariate analysis, TnI >6ng/L compared to <3ng/L was associated with the presence of stenosis, high-risk plaque, and the individual risk features; low attenuation, positive remodeling and spotty calcification. The presence of stenosis was associated with CRP >2mg/L compared to <1mg/L and on a per-risk feature basis, CRP >2mg/L was only associated with the presence of napkin ring sign.

**Table 1:**
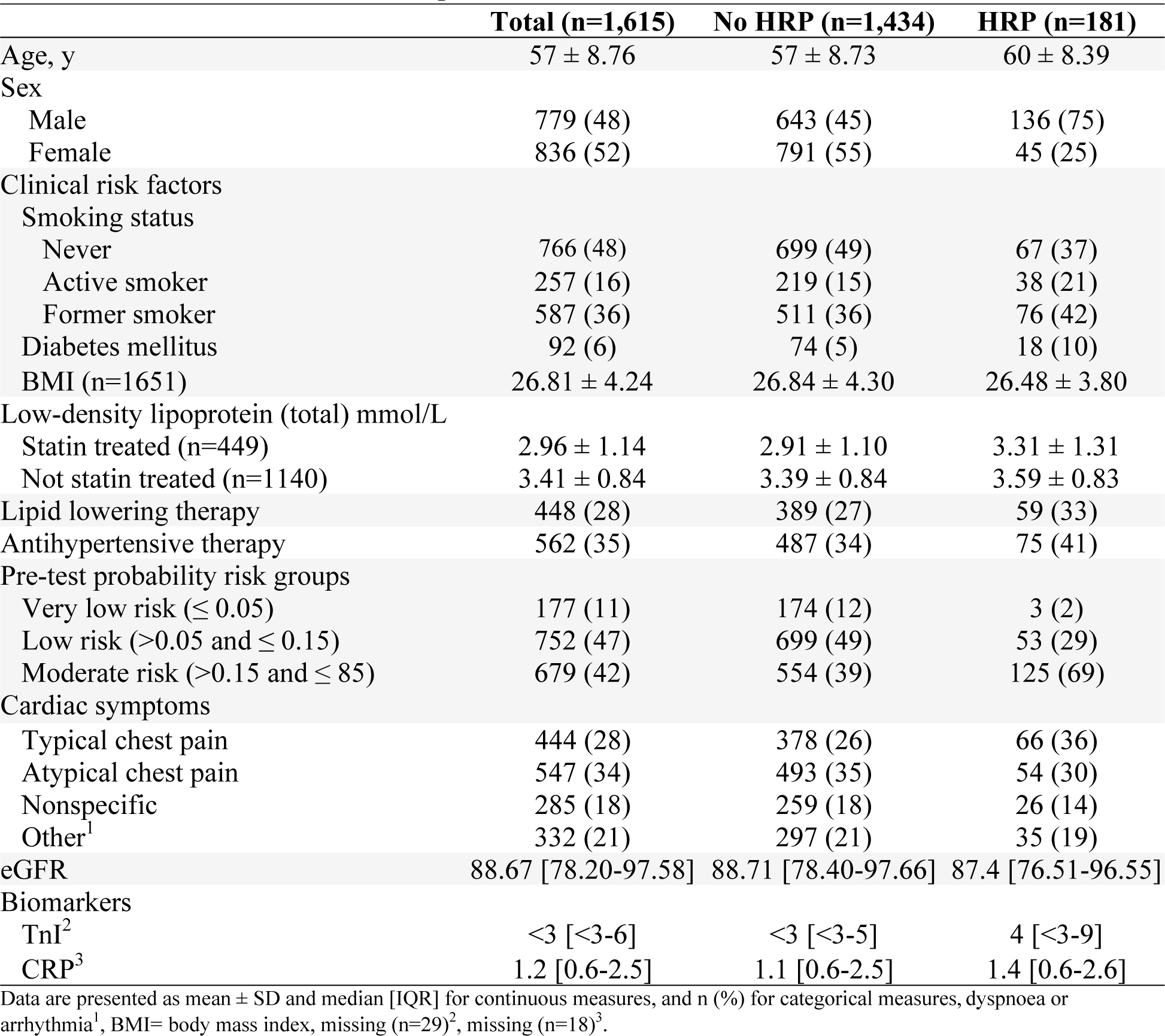
Baseline characteristics in patients with and without HRP.

|  | **Total (n=1,615)** | **No HRP (n=1,434)** | **HRP (n=181)** |
| --- | --- | --- | --- |
| Age, y | 57 ± 8.76 | 57 ± 8.73 | 60 ± 8.39 |
| Sex |  |  |  |
| Male | 779 (48) | 643 (45) | 136 (75) |
| Female | 836 (52) | 791 (55) | 45 (25) |
| Clinical risk factors |  |  |  |
| Smoking status |  |  |  |
| Never | 766 (48) | 699 (49) | 67 (37) |
| Active smoker | 257 (16) | 219 (15) | 38 (21) |
| Former smoker | 587 (36) | 511 (36) | 76 (42) |
| Diabetes mellitus | 92 (6) | 74 (5) | 18 (10) |
| BMI (n=1651) | 26.81 ± 4.24 | 26.84 ± 4.30 | 26.48 ± 3.80 |
| Low-density lipoprotein (total) mmol/L |  |  |  |
| Statin treated (n=449) | 2.96 ± 1.14 | 2.91 ± 1.10 | 3.31 ± 1.31 |
| Not statin treated (n=1140) | 3.41 ± 0.84 | 3.39 ± 0.84 | 3.59 ± 0.83 |
| Lipid lowering therapy | 448 (28) | 389 (27) | 59 (33) |
| Antihypertensive therapy | 562 (35) | 487 (34) | 75 (41) |
| Pre-test probability risk groups |  |  |  |
| Very low risk ($\leq 0.05$) | 177 (11) | 174 (12) | 3 (2) |
| Low risk ($>0.05$ and $\leq 0.15$) | 752 (47) | 699 (49) | 53 (29) |
| Moderate risk ($>0.15$ and $\leq 85$) | 679 (42) | 554 (39) | 125 (69) |
| Cardiac symptoms |  |  |  |
| Typical chest pain | 444 (28) | 378 (26) | 66 (36) |
| Atypical chest pain | 547 (34) | 493 (35) | 54 (30) |
| Nonspecific | 285 (18) | 259 (18) | 26 (14) |
| Other<sup>1</sup> | 332 (21) | 297 (21) | 35 (19) |
| eGFR | 88.67 [78.20-97.58] | 88.71 [78.40-97.66] | 87.4 [76.51-96.55] |
| Biomarkers |  |  |  |
| TnI<sup>2</sup> | <3 [<3-6] | <3 [<3-5] | 4 [<3-9] |
| CRP<sup>3</sup> | 1.2 [0.6-2.5] | 1.1 [0.6-2.5] | 1.4 [0.6-2.6] |

Data are presented as mean ± SD and median [IQR] for continuous measures, and n (%) for categorical measures, dyspnoea or arrhythmia<sup>1</sup>, BMI= body mass index, missing (n=29)<sup>2</sup>, missing (n=18)<sup>3</sup>.

### High-sensitivity Troponin I

Troponin I was measured in 1,604 patients (99.3%), and median TnI was <3 ng/l. Low, moderate, and high TnI levels were observed in 816 (50.9%), 428 (26.7%), and 360 (22.4%) patients, respectively *(supplemental figure 1).* Patients with moderate and high TnI levels were more likely to be men and older than patients with low TnI *(supplemental table 3)*.

#### Association of high-sensitivity Troponin I with plaque burden and stenosis

Troponin I correlated positively but weakly with total plaque burden and plaque burden subtypes; total plaque burden r_s_=0.14, p<0.001; total non-calcified plaque burden r_s_=0.14, p<0.001; total necrotic core plaque burden r_s_=0.13, p <0.001, and CACS r_s_ = 0.15, p <0.001 *(supplemental table 4).* Moreover, the presence of a high total plaque burden and stenosis were both associated with higher TnI levels (p<0.001 for all comparisons) *(figure 3a)*.

**Figure 3:**
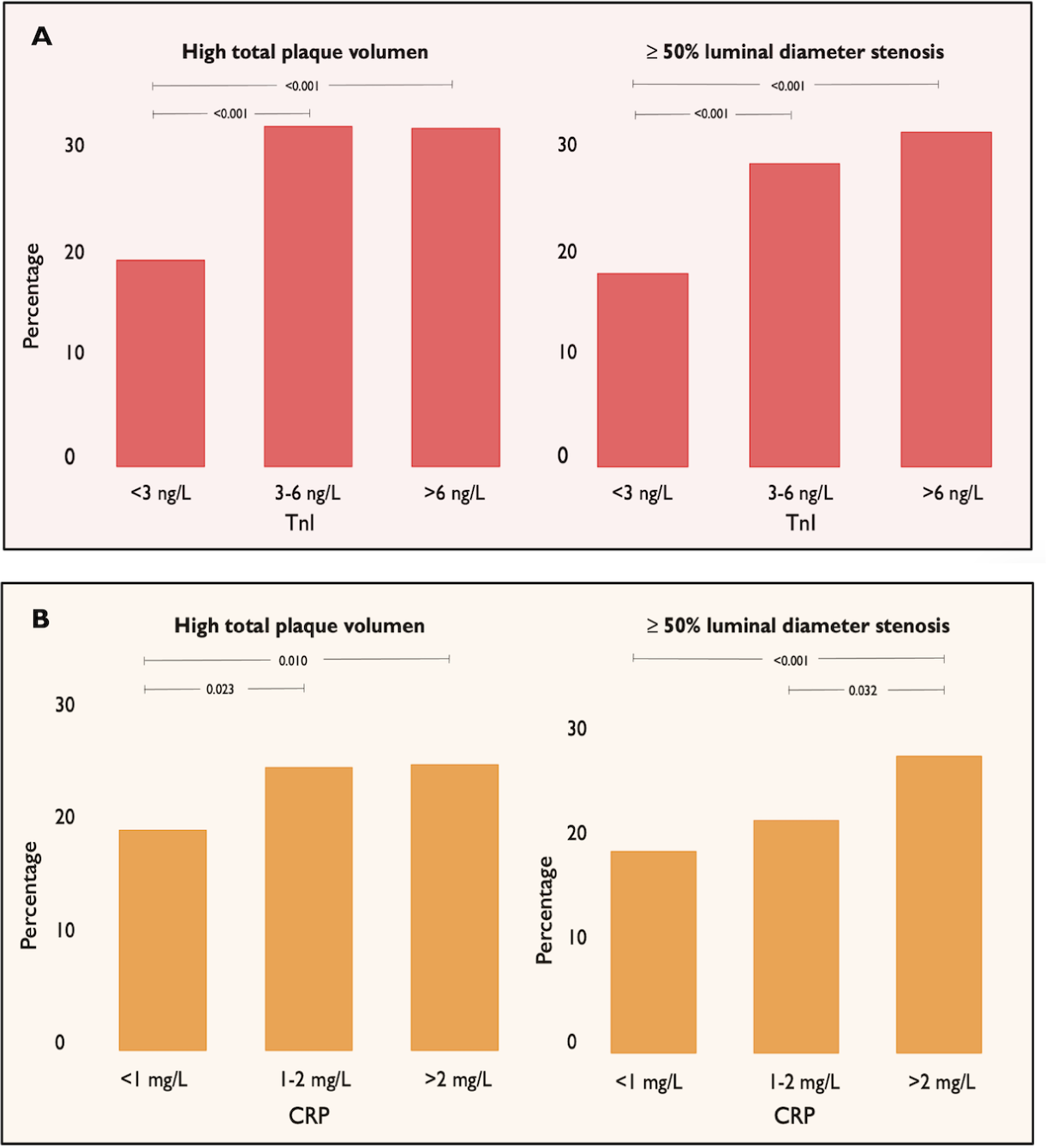
High total plaque burden and stenosis prevalence with increasing TnI (a) and CRP (b) levels. High total plaque burden were defined as total plaque burden >75th percentile, Stenosis was defined as ≥ 50% luminal diameter stenosis.

In multivariate analyses, high TnI was independently associated with the presence of stenosis (OR 1.43, 95% CI: 1.03-1.99, p=0.034) but not high total plaque burden (OR 0.94, 95% CI: 0.69-1.28, p=0.690) *(table 2)*.

**Table 2:**
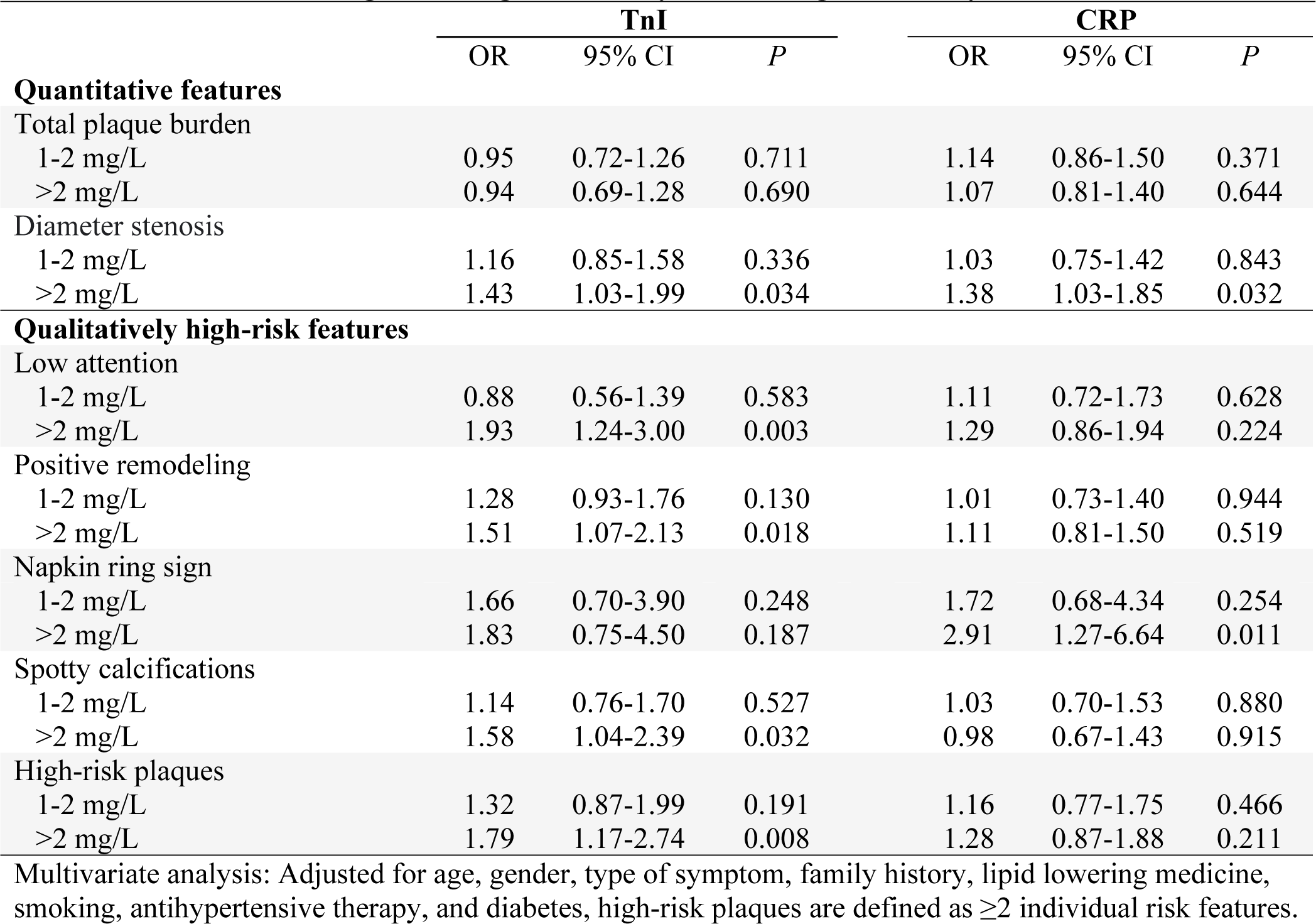
Multivariate regression high-sensitivity TnI and high-sensitivity CRP.

#### Association of high-sensitivity Troponin I with high-risk plaques

Overall, the proportion of patients with HRP increased with rising TnI levels; 60/816 (7.4%) of patients with low TnI had HRP, 58/428 (13.5%) of patients with moderate TnI had HRP, and 62/360 (17.2%) of patients with high TnI had HRP (p = <0.001). Similar findings were present for all individual risk feature except for napkin ring sign *(figure 4a)*.

**Figure 4:**
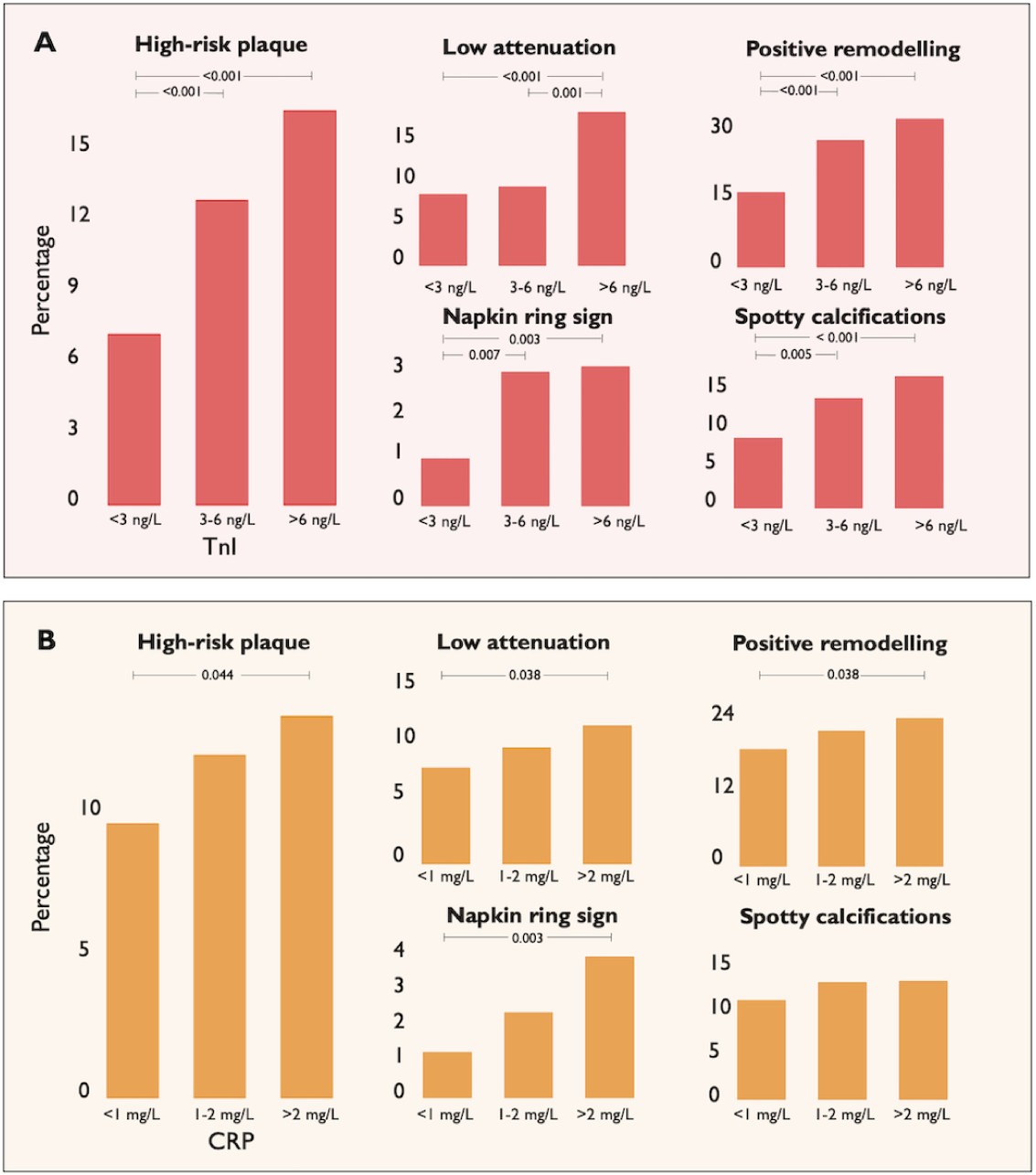
High-risk plaque and individual risk feature prevalence with increasing TnI (a) and CRP (b) levels. A high-risk plaque was defined as a plaque with at least two of the individual risk features.

In univariate analyses, patients with moderate and high TnI levels were at higher risk of having a HRP compared to low TnI level patients. Overall odds for having any individual risk feature were higher in moderate and high TnI groups *(supplemental table 5).* In multivariate analyses, high TnI was associated with having HRP (OR 1.79, 95% CI: 1.17– 2.74, p = 0.008) *(table 2).* For the individual risk features, high TnI levels were associated with low attenuation plaque, positive remodeling, and spotty calcification, however napkin ring sign was not *(table 2)*.

### High-sensitivity C-reactive protein

C-reactive protein was measured in 1,615 patients (100%), and median CRP was 1.2 mg/l. Overall, low, moderate, and high CRP was present in 698/1615 (43.2%), 415/1615 (25.7%), and 502/1615 (31.1%) patients, respectively (supplemental figure 2). Similar to TnI, patients with moderate and high CRP levels were more likely to be men and were older than those with low CRP (supplemental table 6).

#### Association of high-sensitivity C-reactive protein with plaque burden and stenosis

C-reactive protein correlated positively but weakly with total plaque burden and plaque burden subtypes: total plaque burden (r_s_ =0.08, p<0.001); total non-calcified plaque burden (r_s_=0.09, p<0.001); total necrotic core plaque burden (r_s_=0.07, p<0.001); and CACS (r_s_=0.07, p <0.001) *(supplemental table 4).* Moreover, the presence of a high total PAV and stenosis were both associated with increasing CRP levels (p<0.05 for all comparisons) *(figure 3b)*.

In multivariate analyses, high CRP levels were associated with stenosis (OR 1.38, 95% CI: 1.03-1.85, p=0.032) but not high total plaque burden (OR 1.07, 95% CI: 0.81-1.40, p=0.644) *(table 2)*.

#### Association of high-sensitivity C-reactive protein with high-risk plaques

Overall, the proportion of patients having HRPs increased with increasing CRP levels; 66/698 (9.5%) of patients with low CRP having HRP, 49/415 (11.8%) of patients with moderate CRP having HRP, and 66/502 (13.2%) of patients with high CRP having HRP (p=0.044). Similar findings were present of a per HRP-feature basis except for spotty calcification *(figure 4b)*.

In univariate analyses, high CRP levels were associated with HRP *(supplemental table 5).* However, in multivariate analyses, high CRP levels were not associated with the presence of HRP. For the individual high-risk features, high CRP levels were associated with presence of napkin ring sign alone *(table 2)*.

## DISCUSSION

This study is the first of its size to investigate both qualitative and quantitative measures of CAD, assessed as plaque burden and specific plaque features on CCTA, in relation to TnI and CRP. In patients with symptoms suggestive of suspected CAD referred for a primary investigation by CCTA, we found TnI to be associated with the presence of stenosis and HRP, mainly based on an increased prevalence of low attenuation plaques, positive remodeling, and spotty calcification in patients with higher ranges of TnI. Furthermore, we found that CRP was associated with the presence of stenosis and the napkin ring sign feature. Interestingly, high total plaque burden was not linked to TnI and CRP. Our findings indicate that biomarkers of myocardial ischemia and inflammation may be important indicators for identifying patients with stenosis and HRP to improve prognosis.

### Association of high-sensitivity Troponin I with plaque burden and diameter stenosis

Overall, we found no association between elevated TnI and high total plaque burden, and previous studies are ambiguous on the association.^*15*, *24*–*26*^

In line with our findings, the Prospective Multicenter Imaging Study for Evaluation of Chest Pain study (PROMISE) investigators reported an association between elevated levels of TnI and the presence of stenosis. However, their thresholds for high TnI (>1.5 ng/L) were lower compared to our criteria (6 ng/L) which could be due to different detection limits (0.5 ng/L vs. 3.0 ng/L). Similar to our findings, the PROMISE investigators evaluated stenosis as ≥50% diameter stenosis on CCTA which generally overestimates stenosis severity compared to invasive assessment.^*27*^ However, studies in high-risk cohorts with invasive coronary angiography report a similar association between elevated levels of TnI and presence of stenosis.^*28*^

### Association of high-sensitivity Troponin with high-risk plaques and individual risk features

Our findings support the PROMISE trial’s evidence of a positive association between increasing TnI levels and specific HRP features, namely the presence of low attenuation and positive remodeling.^*12*^ In addition, we found TnI levels associated with the presence of spotty calcification which the PROMISE investigators did not include in the HRP feature definition. The PROMISE investigators defined presence of HRP as having just one HRP feature on qualitative CCTA assessment which would improve the ability of TnI to discriminate patients with HRP compared to our definition.

Despite the differences outlined, we similarly found no association between TnI and the presence of napkin ring signs, suggesting that especially low attenuation features and positive remodeling seem associated with elevated TnI levels which previously was confirmed by a small study (N=99) using intravascular ultrasound.^*15*^

### Association of high-sensitivity C-reactive protein with plaque burden and diameter stenosis

Consistent with prior research, other studies reported proportional increasing CRP-levels with increasing diameter stenosis, supporting inflammatory biomarkers as possible predictors of obstructive CAD.^*29*, *30*^ Notably, we found no association between CRP and high total plaque burden *(table 2)* which other authors also confirm.^*12*, *24*, *26*^ Yet, others have associated CRP with specifically non-calcified plaque burden.^*24*^ Overall, whether a link between inflammatory markers and potentially obstructive CAD is caused by systemic inflammation or low-grade ischemia remains unknown. As anti-inflammatory agents successfully decrease the risk of CAD,^*11*, *31*^ a theory of inflammatory markers being important elements in CAD genesis is plausible, and CRP cannot be dismissed when considering CAD extent.

### Association of high-sensitivity C-reactive protein with high-risk plaques and individual risk features

We found no association in multivariate analyses between high CRP and the presence of HRP. On a single-feature basis, only the presence of napkin ring sign was associated with CRP. These results differ from the association reported by the CAPIRE authors between CRP and the presence of low attenuation and positive remodeling.^*26*^ This could partly be explained by differences in populations and improved ability to adjust for confounders based on larger sample size in our multiple regression analysis. Additionally, the CAPIRE investigator did not associate CRP and the presence of the napkin ring sign which can be explained by a reported low feature prevalence and a rather labile definition of low attenuation defined the presence of any voxel <30 HU.^*26*^

The PROMISE investigators found no association between IL-6 and the presence of HRP which is similar to CRP in our study.^*12*^

### Clinical implications

Using CCTA, our findings direct the focus toward plaque characteristics rather than just overall plaque burden, outlining that the presence of stenosis and specifically HRPs may be more important in CAD risk evaluation than the amount of atherosclerosis alone. Our findings suggest that biomarkers can help identify patients with HRP features, which previously was shown to increase risk of future events.^*3*, *32*^ An ongoing prospective cohort, Troponin in Acute Chest Pain to Risk Stratify and Guide Effective Use of Computed Tomography Coronary Angiography (TARGET-CTCA trial; NCT03952351), investigates if patient outcomes are improved by offering CCTA to acute chest pain patients with elevated TnI levels after myocardial infarction is rule out. Likewise, TnI may have a place in pre-test evaluation of patients with stable chest pain by introducing biomarkers to a pre-test clinical likelihood model.

As lipid-lowering medicine has been associated with reduced progression of HRP features^*33*^, offering CCTA to patients with elevated TnI levels in stable chest pain patients may play an important role as it is unknown if all patients with elevated TnI or CRP will benefit from preventive therapy or only those with HRP or other adverse CAD features, Still, trials on biomarker-guided diagnostic testing and medical therapy in de novo stable chest pain patients are warranted.

Large clinical trials showed that anti-inflammatory therapy reduced the rate of recurrent cardiovascular events in patients with CRP ≥2mg/L.^*11*^ The fact that anti-inflammatory therapy reduces cardiovascular events indicates that CRP levels could guide clinicians to introduce anti-inflammatory therapy as a preventive measure for patients with CAD. However, clinical trials testing anti-inflammatory therapy in de novo chest pain patients are warranted.

### Strengths and limitations

The study had limitations. Firstly, categorical data was used as opposed to continuous data and no definite cut-off values for TnI and CRP for identifying stenosis and plaque risk features have been proposed. Our approach validates suggested thresholds from previous studies,^*11*, *15*, *16*^ and the stratification represents a pragmatic use of cut-offs which is helpful in clinical decision-making and structured risk stratification tools. Secondly, a large proportion of patients had undetectable biomarker levels despite highly sensitive assays. A more sensitive analysis could have improved accuracy, but the current approach used commercial assays and clinically relevant cut-offs for real-world relevance. Thirdly, while CRP is the most utilized inflammatory marker in clinical practice, it is also organ-nonspecific; at baseline sampling, reporting on signs of infection might have reduced the risk of competing reasons for elevated CRP. Fourthly, assessment of plaque burden, stenosis, and HRPs is observer-dependent but the applied approach has been validated against intravascular ultrasound and has shown good inter- and intra-observer agreement and reproducibility.^*17*, *34*, *35*^ Additionally, a blinded expert-reader reviewed all lesions with any risk feature to ensure consistency with the pre-defined criteria. Fifthly, we did not have the opportunity to investigate long term outcomes which could have demonstrated the stipulated risk of cardiovascular events for patients with HRPs and whether elevations in biomarkers independent of plaque risk features can predict cardiovascular events. Finally, our population consisted of symptomatic patients referred to non-invasive CCTA and results are only applicable in patients with low and medium risk of CAD and not the general asymptomatic population or in patients with very high-risk.

## CONCLUSION

In de novo chest pain patients, TnI is associated with the presence of stenosis, high-risk plaque, and the individual risk features low attenuation, positive remodeling, and spotty calcification. The presence of stenosis was associated with CRP, and on a per-risk feature basis CRP was only associated with the presence of the napkin ring sign.

## Data Availability

The data that support the results of this study are available from the corresponding author upon reasonable request.

## Abbreviations

CAD: Coronary artery disease
CCTA: Coronary computed tomography angiography HRP High-risk plaque
TnI: Troponin I
CRP: C-reactive protein
FFR: Fractional flow reserve
CACS: Coronary artery calcium score
HU: Hounsfield units
PAV: Percentage atheroma volume

## ACKNOWLEDGEMENTS

The authors acknowledge all the trial site staff, the investigators, and patients who participated in the trial.

## SUPPLEMENTAL MATERIAL

Figures S1-S2.

Tables S1–S6.

